# Effectiveness of Video Teletherapy in Treating Child and Adolescent Obsessive-Compulsive Disorder with Exposure and Response Prevention: a Retrospective Longitudinal Observational Study

**DOI:** 10.1101/2024.07.04.24308232

**Authors:** Jamie D. Feusner, Nicholas R. Farrell, Mia C. Nuñez, Nicholas Lume, Catherine W. MacDonald, Patrick B. McGrath, Larry Trusky, Stephen Smith, Andreas Rhode

## Abstract

**Background:** An effective primary treatment for obsessive-compulsive disorder (OCD) in children and adolescents as well as adults is exposure and response prevention, a form of cognitive-behavioral therapy. Despite strong evidence supporting the efficacy and effectiveness of exposure and response prevention from studies in research and real-world settings, its clinical use remains limited. This underuse is often attributed to access barriers such as the scarcity of properly trained therapists, geographical constraints, and costs. Some of these barriers may be addressed with virtual behavioral health, providing exposure and response prevention for OCD through video teletherapy and supplemented by app-based therapeutic tools and messaging support between sessions. While studies of teletherapy exposure and response prevention in adults with OCD have shown benefits in research and real-world settings in both small and large samples, studies in children and adolescents thus far have been in small samples and limited to research settings.

**Objective:** This study reports on the real-world effectiveness of teletherapy exposure and response prevention for OCD in the largest sample (N=2173) of child and adolescent patients to date.

**Methods:** Children and adolescents with OCD were treated with live, face-to-face video teletherapy sessions, with parent/caregiver involvement, using exposure and response prevention. Assessments were conducted at baseline, after 7-11 weeks, and after 13-17 weeks. Additionally, longitudinal assessments of OCD symptoms were performed at weeks 18-30, 31-42, and 43-54.

**Results:** Treatment resulted in a median 37.3% decrease in OCD symptoms at 13-17 weeks, and 53.4% of youth met full response criteria at this point. In addition, there were significant reductions in the severity of depression, anxiety, and stress symptoms. The median amount of therapist involvement was 13 appointments and 11.5 hours. Further, symptom improvements were maintained, or improved upon, in the longitudinal assessment period.

**Conclusions:** These results show that remote ERP treatment, assisted by technology, can effectively improve symptoms in children and adolescents with OCD in a real-world setting. Notable outcomes were achieved in a relatively small amount of therapist time, demonstrating its efficiency. These findings have implications for widespread dissemination of accessible, evidence-based care for children and adolescents with OCD.

## Introduction

Obsessive-compulsive disorder (OCD) is a common and often disabling mental health condition that affects 0.25-4% of children and adolescents.^1–3^ Without treatment, OCD can persist into adulthood. Further, it can significantly interfere with a young person’s development, education, and relationships. Fortunately, OCD in young people can be effectively managed through psychotherapy, medication, or a combination of the two. Exposure and response prevention (ERP), a form of cognitive-behavioral therapy (CBT), is particularly effective for treating OCD in children and adolescents. It has been extensively tested in clinical trials^4–6^ and is recommended as the first-line treatment for OCD.^4,7–11^

However, accessing ERP can be challenging due to a shortage of therapists trained in this specialized technique, along with the costs and geographical limitations of attending in-person therapy sessions. Therapists with adequate training in ERP for children and adolescents are even more scarce.^12,13^ Recognizing these obstacles, NOCD has created a virtual therapy program that delivers ERP through video teletherapy. Remote video telemedicine treatment, in general, has shown non-inferiority to traditional in-person treatment for OCD, anxiety, and depression in a head-to-head study in adults.^14^ Meta-analyses of remote delivery, specifically, of CBT for OCD have shown benefits for adults and children/adolescents.^15,16^ Non-inferiority of remote CBT delivered by telephone compared with face-to-face CBT has also been demonstrated in adolescents with OCD.^17^ Remote treatments have the added benefit of allowing therapy to take place in the patient’s everyday environment. This can be especially helpful for younger patients, as it enables therapists to work with them in the settings that trigger their OCD symptoms, such as at home or school.

The convenience of teletherapy, coupled with the widespread ownership of smartphones, makes this form of treatment a promising option for families seeking help for OCD. NOCD has created a virtual therapy program that delivers ERP through video teletherapy, aiming to maximize therapeutic impact while minimizing the time required from therapists. The program uses treatment design elements from an open clinical trial in adults that demonstrated the effectiveness of combining the NOCD app with face-to-face therapy sessions in reducing OCD symptoms significantly and efficiently, while achieving good patient satisfaction.^18^ To extend the reach of its treatment and enhance its effectiveness, NOCD provides remote sessions via video teletherapy, along with additional support through messaging with a therapist, an online OCD community, and peer support. We previously reported clinical outcomes from N=3552 adults with OCD treated with ERP.^19^ In this retrospective observational study, the median improvement was a 45% reduction in OCD symptoms. Further, 62.9% met the criteria for full response.

The treatment for children and adolescents at NOCD follows a similar structure as for adults, although it includes involvement of parents and/or other caregivers. This comprehensive model aims to improve access to effective OCD treatment for children and adolescents, thereby addressing a significant gap in mental health services for young people. The goal of the current retrospective observational longitudinal study was to determine clinical outcomes in a large naturalistic sample of children and adolescents with a primary diagnosis of OCD who received ERP treatment via video teletherapy.

## Methods

### Initial Evaluation and Clinical Assessments

As part of the general clinical enrollment at NOCD, parents or legal guardians initially reached out to the intake team as self-referrals or received a referral from their insurance or medical provider. Therapists trained by NOCD in OCD assessment and treatment conducted the initial diagnostic evaluations. These evaluations occurred over the first two sessions and included a thorough clinical review covering the biopsychosocial aspects of the individual’s history and a semi-structured diagnostic interview using the Diagnostic Interview for Anxiety, Mood, and Obsessive-Compulsive and Related Neuropsychiatric Disorders (DIAMOND).^20^ Those diagnosed with OCD as their primary concern, according to DSM-5 criteria^21^ and the DIAMOND, received treatment. The majority of candidates with an “extreme” rating on the DIAMOND clinician-rated severity scale were directed to more intensive treatment options, although exceptions were made based on clinician judgment. Referrals for other significant psychiatric or substance use issues, if they were deemed to potentially interfere with ERP treatment, were also made as needed (e.g. to child and adolescent psychiatrists or other specialty providers). NOCD generally provides services to individuals aged 5 and above, although some exceptions were made for some 4-year-old patients.

### Treatment Approach

The treatment plan included weekly or twice-weekly 60-minute ERP sessions via video for the first three weeks for most, followed by, typically, 10–14 weeks of weekly 60-minute sessions to support the continuation of ERP exercises. During this phase, some transitioned to 30-minute check-in sessions based on their clinical progress. Therapists, while aiming to adhere to this structured approach, could adjust the number or frequency of sessions to meet clinical needs and/or accommodate patients’ and their parents’ schedules. Family therapy sessions were scheduled when the therapist deemed them necessary. Additionally, patients and their parents could engage in asynchronous in-app messaging with their therapists for guidance on homework assignments or for support. NOCD also offered round-the-clock support through online monitored community groups. Patients and their parents also had access to the NOCD app for tools to set up ERP exercises with their therapist, record the results of their ERP exercises, and to access and interact with the online NOCD community. Parents and caregivers of youth with OCD were able to access weekly support groups facilitated by a trained clinician to receive ongoing support and guidance in reducing accommodation of OCD in the home. Lastly, parents and caregivers received personalized psychoeducation on OCD symptoms in youth and on the maintaining role of familial accommodation, and were given guidance in developing a plan to progressively reduce and eventually eliminate any accommodation behaviors.

All sessions utilized a secure, U.S. Health Insurance Portability and Accountability Act (HIPAA)-compliant (and compliant with other country’s health information privacy regulations) version of Zoom, accessible via personal computing or mobile devices, with live technical support available during business hours to address connectivity issues.

### Therapist Qualifications and Training

Study therapists held Master’s or doctoral degrees and were licensed in the states of patients to whom they provided treatment or were associate clinicians who were supervised by licensed therapists. Therapists underwent comprehensive ERP training from NOCD. This included multiple days of focused instruction on OCD and ERP techniques. This was followed by evaluations including written tests and mock diagnostic, education, and ERP treatment sessions. Therapists were required to pass these written and practical evaluations before starting to treat patients. Additionally, clinicians received further training and guided practice opportunities in the following areas: (a) providing psychoeducation to youth using age-appropriate language and examples, (b) tailoring ERP for OCD in youth, and (c) addressing familial accommodation. Ongoing consultation was provided, including weekly group sessions and periodic case reviews.

### Assessments

Patients completed self-report assessments to avoid therapist bias. These included the Dimensional Obsessive-Compulsive Scale (DOCS), which was the primary OCD outcome measure, and the Depression, Anxiety, and Stress Scales (DASS-21) to measure commonly occurring comorbid depressive, anxiety, and stress symptoms. Links to these assessments were sent to patients/parents via the NOCD app every three weeks. Therapists also completed the DIAMOND severity scale, a clinician-rated measure of OCD severity.

The Dimensional Obsessive-Compulsive Scale (DOCS)^22^ is a 20-item self-report measure of OCD symptom severity across four domains: contamination, responsibility for harm or mistakes, unacceptable thoughts, and incompleteness/symmetry. The DOCS has shown good psychometric properties, including strong convergent validity with the Yale-Brown Obsessive Compulsive Scale (r =.54) and the Obsessive-Compulsive Inventory-Revised (r =.69), and is sensitive to the effects of treatment. Further, it shows strong correlations with the Obsessive-Compulsive Inventory-Children’s Version Revised^23^ at baseline (r=.77), at 3 weeks (r=.80), and at 9 weeks (r=.80) of treatment.^24^

The Diagnostic Interview for Anxiety, Mood, and Obsessive Compulsive and Related Neuropsychiatric Disorders (DIAMOND) severity scale^20^ is a 2-item clinician-rated assessment of the overall severity of an individual’s emotional distress and functional impairment related to OCD symptoms. The clinician makes separate ratings of an individual’s emotional distress and functional impairment on a scale ranging from 1 (Normal) to 7 (Extreme), and the higher of the two ratings is taken as the total severity score.

The Depression, Anxiety, and Stress Scales (DASS-21)^25^ is a 21-item self-report measure of symptoms of depression, anxiety, and stress. It has been widely used in previous research and has consistently shown good psychometric qualities.

### Statistical Analysis

Data, anonymized before analysis, were examined using a linear mixed model approach, with time points as fixed factors and patients as random factors. The time points for rating scale scores used in the linear mixed model analysis included ratings at baseline, the most recent rating obtained between weeks 7-11, and the most recent rating obtained between weeks 13-17. Since there was some degree of flexibility in the treatment, not everyone had rating scales done at precisely the same session or week of their treatment. Thus, these windows allowed us to measure symptom improvement at approximately a midpoint in treatment (weeks 7-11) and at the end of the active treatment period (weeks 13-17). The primary (DOCS scores) and secondary (DASS-21 depression, anxiety, and stress scores) outcome measures were analyzed accordingly, with statistical significance set at an alpha of 0.05. We calculated effect sizes using Hedges’ *g*. Descriptive statistics, including treatment duration and mean and median symptom improvements, were calculated for those who had a baseline and at least one subsequent rating at 13–17 weeks. Analyses were conducted in R.

### Ethical Considerations

The analyses in this study did not require research ethics board review as this does not meet the criteria for Human Subject Research as defined by federal regulations for human subject protections, 45 CFR 46.102(e); this is a secondary analysis of de-identified data from clinical records, obtained and analyzed retrospectively, and was not the result of a research intervention or interaction. Compliance with data protection laws was ensured through NOCD’s privacy policy, which all patients agreed to, outlining data use and protection measures.

## Results

### Sample

We analyzed data, retrospectively, from children and adolescents with a primary obsessive-compulsive disorder diagnosis who received ERP treatment at NOCD from Jul 23, 2020 (when NOCD began offering child and adolescent treatment) to May 3, 2024, when we froze the data for analysis. The total sample size of individuals who had at least a baseline and a week 13-17 DOCS completed was N=2173. The number of individuals who had a baseline, week 7-11, and a week 13-17 DOCS completed was n=1797.

### App use and messaging

In terms of app usage, n=2129 (97.98%) opened the app more than 10 times. N=2127 (97.88%) had at least one chat message with their therapist, while n=1835 (84.45%) had 10 or more chat messages. The mean number of chat messages for those who had at least one message was 53.53±87.

### Treatment duration

The mean treatment duration was 15.12 ± 7.96 weeks (median = 15, interquartile range = 10.86 – 17.00, mode = 16), the mean number of visits was 13.70 ± 5.83 (median = 13, interquartile range = 10.00 – 16.00, mode = 13), and the mean number of therapist hours was 12.60 ± 5.58 (median = 11.50, interquartile range = 9.00 – 15.00, mode = 10.50).

### OCD symptom results

NOCD treatment resulted in a significant decrease in patient-rated OCD symptoms over time (DOCS scores) (effect of time: t_6927.69_=-52.77, *P*<.001; initial to endpoint Hedges g = -.65: “medium” effect size, CI -0.59 to -0.70). From baseline to week 7-11, DOCS scores decreased from a mean of 27.4±14.1 to a mean of 19.7±13.1, representing a mean -7.7 (CI -7.1 to -8.2) point decrease (28.10%). By week 13-17, DOCS scores improved to a mean of 18.3±12.9, representing a mean 9.1 (CI -8.5 to -9.7) point decrease (33.2%) (see Fig. 2 and Table 2). On the individual patient level, median DOCS score improvement was 38.46% (interquartile range = 12.50% to 64.00%). An exploratory analysis of responses in children (ages 4-12) and adolescents (ages 13-17), separately, revealed similar results (children, the effect of time on DOCS scores: β=-5.10, t_2132.88_=-27.06, *P*<.001. In adolescents, the effect of time on DOCS scores: β=-5.39, t_4071.89_=-40.99, *P*<.001. An additional exploratory analysis testing the effects of ethnic/racial categories did not reveal any significant effects of these categories on the overall outcomes (all p values ≥.1).

**Table 1.**
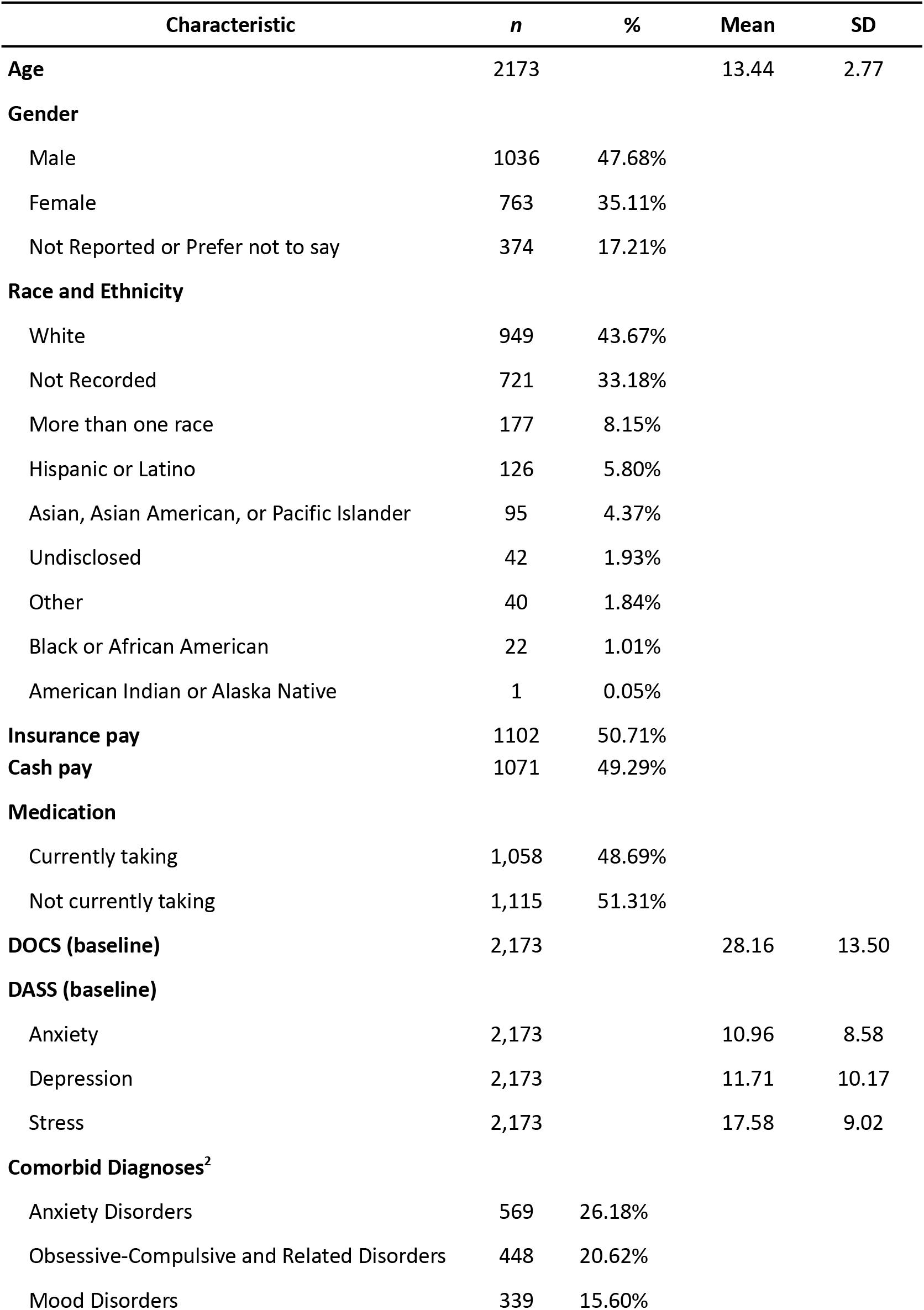
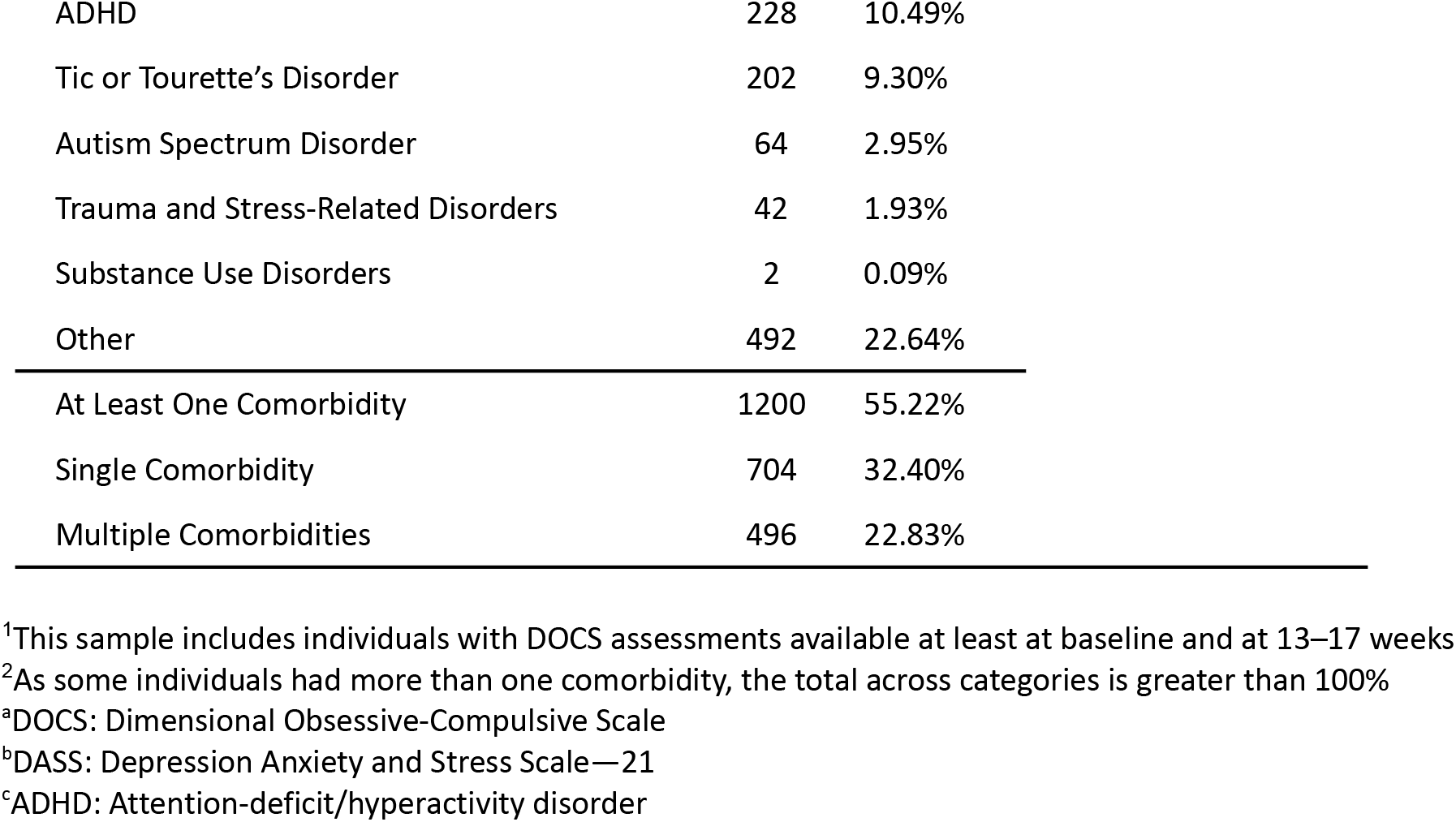
Demographics and psychometrics^1^.

| Characteristic | *n* | % | Mean | SD |
| --- | --- | --- | --- | --- |
| **Age** | 2173 |  | 13.44 | 2.77 |
| **Gender** |  |  |  |  |
| Male | 1036 | 47.68% |  |  |
| Female | 763 | 35.11% |  |  |
| Not Reported or Prefer not to say | 374 | 17.21% |  |  |
| **Race and Ethnicity** |  |  |  |  |
| White | 949 | 43.67% |  |  |
| Not Recorded | 721 | 33.18% |  |  |
| More than one race | 177 | 8.15% |  |  |
| Hispanic or Latino | 126 | 5.80% |  |  |
| Asian, Asian American, or Pacific Islander | 95 | 4.37% |  |  |
| Undisclosed | 42 | 1.93% |  |  |
| Other | 40 | 1.84% |  |  |
| Black or African American | 22 | 1.01% |  |  |
| American Indian or Alaska Native | 1 | 0.05% |  |  |
| **Insurance pay** | 1102 | 50.71% |  |  |
| **Cash pay** | 1071 | 49.29% |  |  |
| **Medication** |  |  |  |  |
| Currently taking | 1,058 | 48.69% |  |  |
| Not currently taking | 1,115 | 51.31% |  |  |
| **DOCS (baseline)** | 2,173 |  | 28.16 | 13.50 |
| **DASS (baseline)** |  |  |  |  |
| Anxiety | 2,173 |  | 10.96 | 8.58 |
| Depression | 2,173 |  | 11.71 | 10.17 |
| Stress | 2,173 |  | 17.58 | 9.02 |
| **Comorbid Diagnoses<sup>2</sup>** |  |  |  |  |
| Anxiety Disorders | 569 | 26.18% |  |  |
| Obsessive-Compulsive and Related Disorders | 448 | 20.62% |  |  |
| Mood Disorders | 339 | 15.60% |  |  |

|  |  |  |
| --- | --- | --- |
| ADHD | 228 | 10.49% |
| Tic or Tourette's Disorder | 202 | 9.30% |
| Autism Spectrum Disorder | 64 | 2.95% |
| Trauma and Stress-Related Disorders | 42 | 1.93% |
| Substance Use Disorders | 2 | 0.09% |
| Other | 492 | 22.64% |
| At Least One Comorbidity | 1200 | 55.22% |
| Single Comorbidity | 704 | 32.40% |
| Multiple Comorbidities | 496 | 22.83% |

<sup>1</sup>This sample includes individuals with DOCS assessments available at least at baseline and at 13–17 weeks
<sup>2</sup>As some individuals had more than one comorbidity, the total across categories is greater than 100%
<sup>a</sup>DOCS: Dimensional Obsessive-Compulsive Scale
<sup>b</sup>DASS: Depression Anxiety and Stress Scale—21
<sup>c</sup>ADHD: Attention-deficit/hyperactivity disorder

**Table 2.** Clinical assessments by treatment time point.

| Outcome scale and assessment time point | Valid, n | Missing<sup>1</sup>, n | Mean | SD |
| --- | --- | --- | --- | --- |
| **DOCS<sup>a</sup>** |  |  |  |  |
| Baseline | 2173 | 0 | 28.16 | 13.50 |
| Week 7-11 | 1797 | 376 | 20.58 | 13.00 |
| Week 13-17 | 2173 | 0 | 17.94 | 12.90 |
| Week 18-30 | 1779 | 394 | 15.26 | 12.01 |
| **Week 31-42** | 1257 | 916 | 14.04 | 11.71 |
| **Week 43-54** | 859 | 1314 | 13.51 | 11.87 |
| **DASS<sup>b</sup> depression** |  |  |  |  |
| **Initial** | 2173 | 0 | 11.71 | 10.17 |
| **Midpoint** | 1527 | 646 | 9.00 | 9.38 |
| **Endpoint** | 1766 | 407 | 8.04 | 9.19 |
| **DASS anxiety** |  |  |  |  |
| **Initial** | 2173 | 0 | 10.96 | 8.58 |
| **Midpoint** | 1527 | 646 | 7.94 | 7.63 |
| **Endpoint** | 1766 | 407 | 6.93 | 7.18 |
| **DASS stress** |  |  |  |  |
| **Initial** | 2173 | 0 | 17.58 | 9.02 |
| **Midpoint** | 1527 | 646 | 13.65 | 8.79 |
| **Endpoint** | 1766 | 407 | 12.60 | 8.73 |

<sup>1</sup>Missing data at week 7-11 for DOCS and DASS scales was due to the patient not completing the rating scale(s). Missing data for DOCS at weeks 18-30, 31-42, or 43-54 was due to either the patient not completing the rating scale or the patient having left treatment
<sup>a</sup>DOCS: Dimensional Obsessive-Compulsive Scale
<sup>b</sup>DASS: Depression Anxiety and Stress Scale—21

**Fig. 1.**
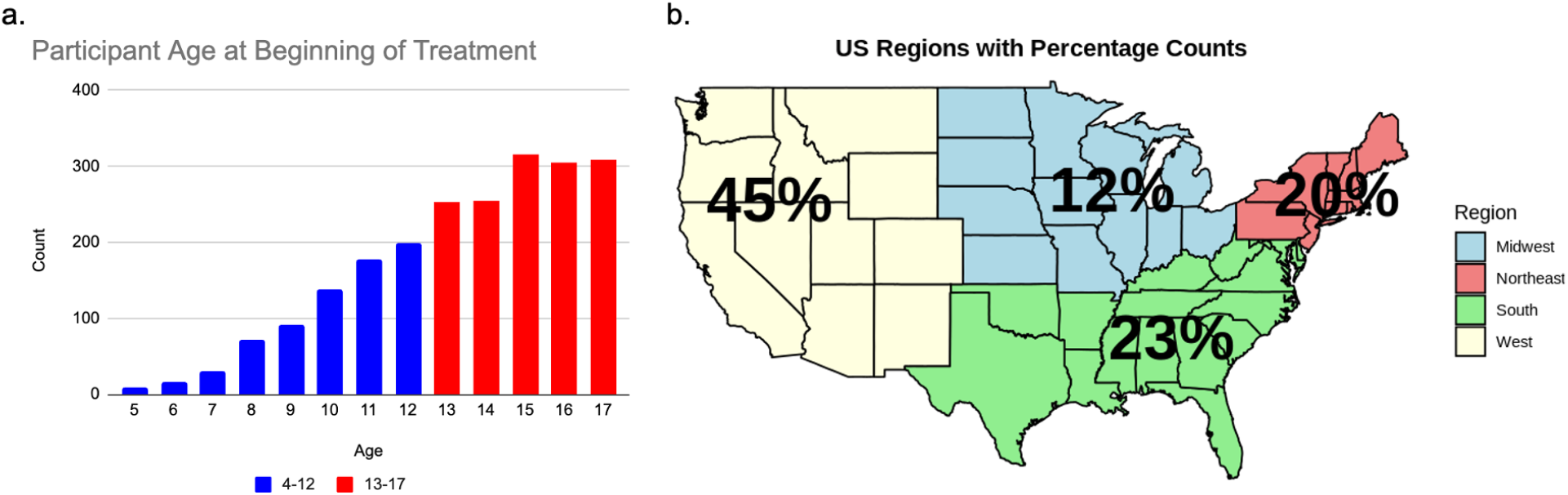
Age and U.S. geographic distributions for the total sample. a. Participant age at the beginning of treatment. Blue indicates children and red indicates adolescents. b. Percentages of patients by U.S. geographic location of residence.

**Figure 2.**
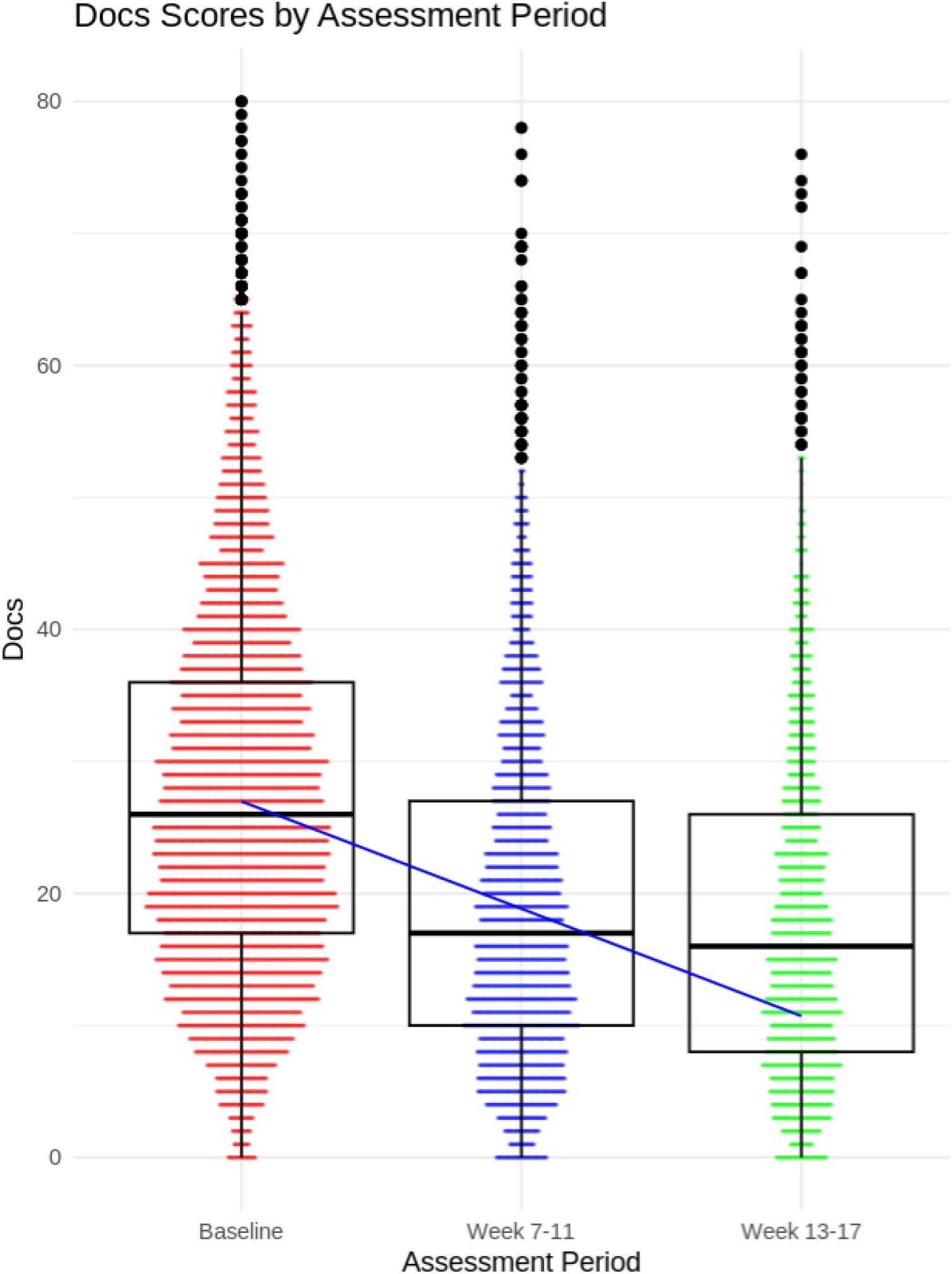
Changes in obsessive-compulsive disorder symptoms as assessed by the Dimensional Obsessive-Compulsive Scale (DOCS) with treatment. Median and interquartile ranges are indicated in the box-and-whisker plots. *P*<.001 for the effect of assessment period.

Further, 53.4% had ≥35% reduction in OCD symptoms and were categorized as full “responders”^26^. A total of 62.0% achieved either partial (25-35% reduction) or full response.

### Depression, Anxiety, and Stress Results

Treatment resulted in significant improvements on the DASS depression (t_6415.03_=-30.55, *P*<.001; initial to endpoint Hedges g = -.37, CI -0.32 to -0.42), DASS anxiety (t_6571.91_=-34.05, *P*<.001; initial to endpoint Hedges g = -.43, CI -0.38 to -0.49), DASS stress (F_7123.66_=-36.66, *P*<.001; initial to endpoint Hedges g = -.52, CI -0.47 to -0.57) (see Fig. 3 and Table 2).

**Figure 3.**
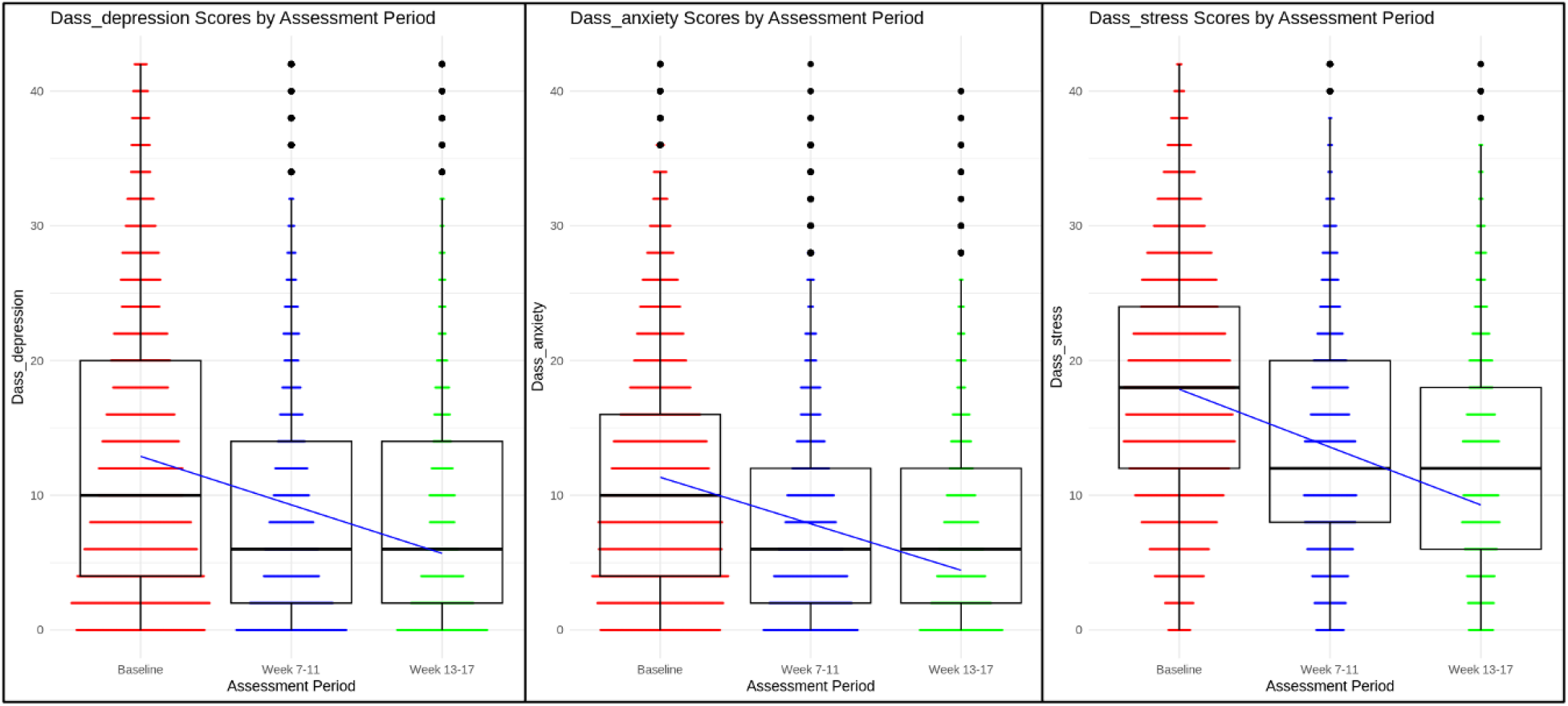
Changes in a) depression, b) anxiety, and c) stress symptoms with treatment, as assessed by the Depression Anxiety and Stress Scale (DASS-21). Median and interquartile ranges are indicated in the box-and-whisker plots. *P*<.001 for the effect of assessment period.

### Post hoc analysis of outcomes stratified by starting clinician-rated severity level

To determine how treatment response differed by different initial severity levels of OCD, we used the DIAMOND clinician-rated severity scale at the initial assessment to stratify patients into three groups of severity ratings: “Mild” (severity score of 2 or 3), “Moderate” (severity score of 4 or 5), or “Severe” (severity score of 6 or 7) (n=4 had missing DIAMOND data). For DOCS scores, on the individual patient level the Mild group (n=264) had a median 40.31% reduction (interquartile range: 8.51%-79.80%), the Moderate group (n=1761) a median 38.36% reduction (interquartile range: 13.33%-63.64%), and the Severe group (n=144) a median 34.07% reduction (interquartile range: 6.56%-58.52%). Response rates from the DOCS were 55.3% for Mild, 54.23% for Moderate, and 49.30% for Severe.

### Longitudinal follow-up

We conducted an analysis of longitudinal follow-up of OCD symptom scores after the active treatment period (weeks 13-17). At the follow-up windows of weeks 18-30, 31-42, and 43-54, most patients maintained their gains or made further improvements (effect of time: t_7970.47_=-61.99, *P*<.001) (see Fig. 4 and Table 2). From baseline to week 18-30 (n=1779), DOCS scores decreased from a mean of 28.16 to a mean of 15.26, representing a mean -12.9 (CI -12.11 to -13.7) point decrease (45.81%), Hedges g = -1, CI = -0.94 to -1.07. By weeks 31-42 (n=1257), DOCS scores improved to a mean of 14.04, representing a mean -14.12 (CI -13.27 to -14.99) point decrease (50.14%), Hedges g = -1.09, CI = -1.02 to -1.17. By weeks 43-54 (n=859), DOCS scores improved to a mean of 13.51, representing a mean -14.65 (CI -13.68 to -15.63) point decrease (52.02%) from baseline, Hedges g = -1.12, CI = -1.04 to -1.2.

**Figure 4.**
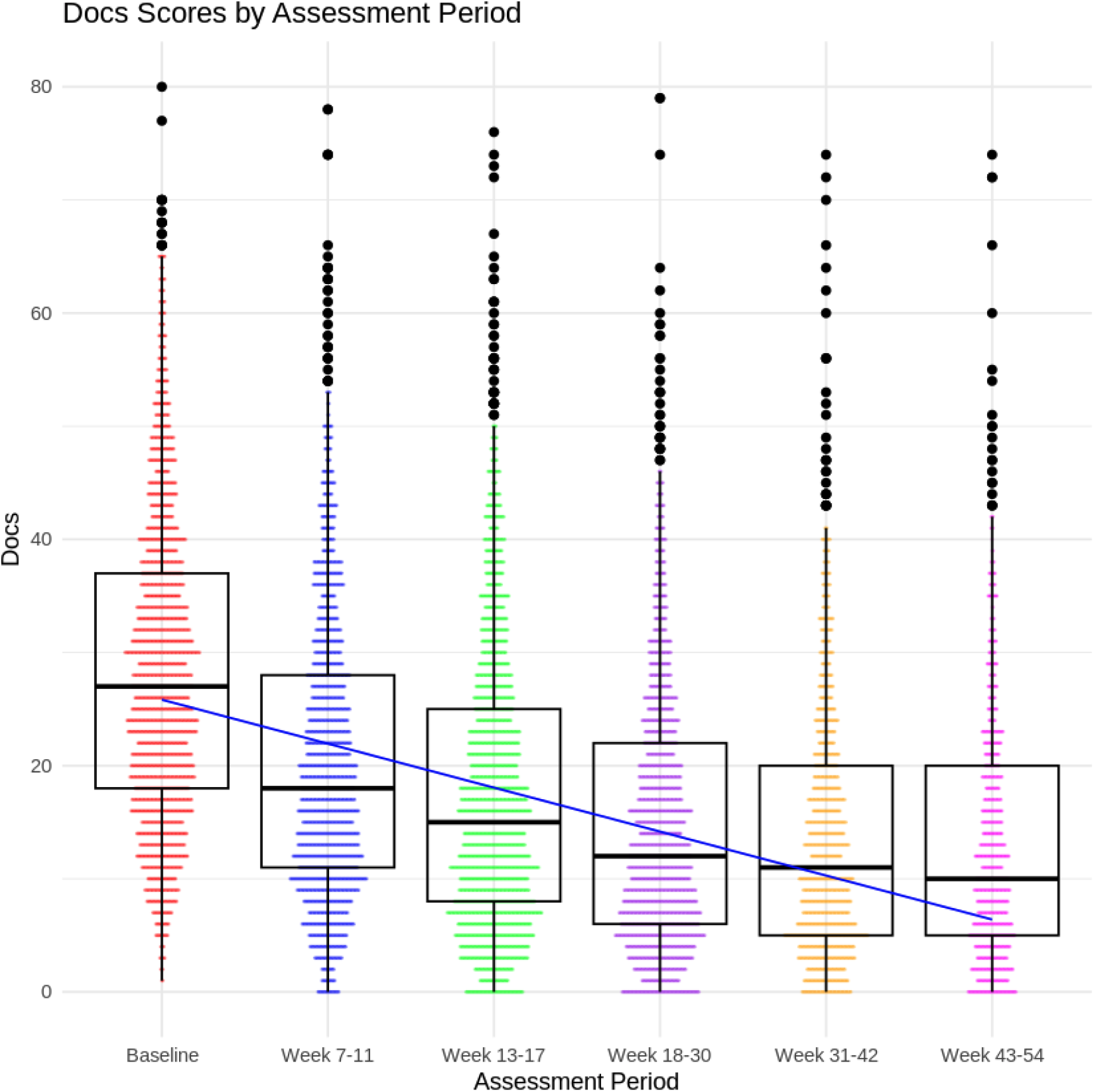
Longitudinal follow-up of obsessive-compulsive disorder symptoms as assessed by the Dimensional Obsessive-Compulsive Scale (DOCS) Median and interquartile ranges are indicated in the box-and-whisker plots. *P*<.001 for the effect of assessment period.

## Discussion

The aim of this retrospective observational longitudinal study was to assess the clinical outcomes of a large, naturalistic sample of children and adolescents with a primary diagnosis of OCD who received ERP treatment using video teletherapy. Children and adolescents exhibited significant reductions in their symptoms, demonstrating the effectiveness of this approach for younger populations. Specifically, symptom reduction was notable with a median 37.3% reduction, 53.4% of participants achieving full response, and 62.0% showing either partial or full response. The treatment also led to improvements in commonly co-occurring symptoms such as depression, anxiety, and stress. There were similar outcomes for children as for adolescents, and outcomes did not significantly differ by ethnic/racial categories. Moreover, the results were largely maintained, or improved upon, for those in long term follow-up. These outcomes underscore the potential of targeted OCD treatment to alleviate a range of severe and distressing symptoms that stem from their primary OCD diathesis. This is particularly important given that an estimated 50-87.5% of OCD cases onset before the age of 21^27,28^ and OCD is a chronic condition if left untreated. Further, individuals often endure symptoms for an average of 11 years before receiving treatment. ^29^

The results highlight the substantial impact and efficiency of this treatment model for OCD and associated symptoms, offering both time and cost savings. The rapid timeframe of these improvements, achieved in a median of 13 sessions and 11.5 hours, represents a significant reduction in both therapist time and treatment duration compared to what has been observed in treatment-as-usual outpatient CBT (37.0 ±45.0 sessions).^30^ This efficiency has implications for considerable cost reductions for families and insurance providers.

The treatment methodology for children and adolescents, as for adults, was influenced by a previously developed and evaluated approach to provide evidence-based ERP treatment efficiently in terms of therapist time.^18^ The symptom reduction achieved in this study aligns with that found in earlier research, although direct comparisons are somewhat limited due to differences in setting (real-world clinical versus controlled research environments) and outcome measures used (patient-rated DOCS versus clinician-rated Yale-Brown Obsessive-Compulsive Scale).^29^ Additionally, the use of face-to-face teletherapy distinguishes this treatment from in-person methods previously studied.

In terms of comparison with previously-reported adult outcomes,^19^ the results in this child and adolescent cohort show a slightly lower magnitude of symptom reduction (median 7.7% lower). This may be due to several factors. Children and adolescents may not as readily comprehend the rationale behind ERP, which might result in some resistance in completing all required homework. In general, the idea of intentionally experiencing distress in the interest of overcoming symptoms may be highly counterintuitive, especially for children. The concepts of habituation with repeated exposures and interruption of compulsions as a way to break the cycle of obsessions and compulsions are abstract concepts that may exceed the cognitive developmental capacities of some. These potential barriers are partially mitigated, however, by parental psychoeducation and their involvement in the treatment.

Further, in this cohort, symptom improvements were relatively consistent across mild, moderate, and severe cases. This indicates the treatment model’s broad applicability and effectiveness across different severity levels of OCD, even in those with severe OCD, who achieved a median of 34.07% symptom reduction. This finding emphasizes the treatment’s capacity to address the needs of a diverse group of young patients in a time-efficient manner.

An innovative aspect of the NOCD model was the inclusion of additional patient support mechanisms, such as between-session SMS messaging with therapists, which was used by about 98% of members and in those who used it at an average of about 54 times. Also available was 24-hour access to NOCD’s online support community, which could facilitate a sense of belonging and understanding among participants and their parents or caregivers but also helped normalize their experiences by connecting them with peers facing similar challenges. This peer support, especially from individuals who had successfully completed the NOCD treatment, likely encouraged ongoing engagement and adherence to the therapy process, which is critical given the inherently challenging nature of ERP.

The use of technology, including video teletherapy and integrated communication tools, was pivotal in engaging and effectively treating a wide demographic of young patients across various locations. These technological solutions allowed for the execution of in-session, in-vivo exercises tailored to the individual’s symptoms and environments, enhancing the relevance and impact of the therapy. Previous research supports the efficacy of remote therapy, and the significant symptom improvement rates observed in this study further validate the effectiveness of virtual ERP, comparable to traditional in-person therapy.

There are limitations to this analysis. These include its observational design and the lack of standardized treatment fidelity checks typically found in controlled trials, although NOCD therapists received the same, standardized training, and were audited for their adherence to their ERP training and their achieved outcomes. Importantly, however, the flexibility allowed within the treatment model provides real-world applicability and potential for adaptation to the highly-varied individual patient and parent needs.

## Conclusions

Overall, ERP delivered via technology-assisted video teletherapy results in clinically significant improvements in OCD, depression, anxiety, and stress symptoms for children and adolescents with OCD. This is achieved in approximately 65% fewer sessions than treatment-as-usual outpatient ERP/CBT. This could translate to lower costs than such lower-intensity approaches, and may also translate to reduced costs compared with higher-intensity programs for those with severe symptoms. Further, it is effective for moderate and severe OCD, which for some may prevent the need for higher levels of care such as intensive outpatient, partial hospitalization, residential, or inpatient treatment. Because OCD in this population intimately involves the family, healthier children and adolescents could translate to less stress and better health in family members. In sum, this treatment modality offers a scalable, effective option for accessing evidence-based care, potentially reducing the burden of OCD on young individuals and their families while also presenting an opportunity for significant cost savings compared to traditional treatment methods.

## Data Availability

All data produced in the present analysis are available upon reasonable request to the authors

## Acknowledgments

The authors would like to thank the NOCD therapists and Member Advocates for their help in facilitating treatment and care experiences.

There was no external funding for the analysis.

## Conflicts of Interest

JDF, NRF, MN, NL, CM, PM, LT, SS, and AR report personal fees from NOCD Inc.

## Abbreviations

OCD: obsessive-compulsive disorder
ERP: exposure and response prevention
DOCS: Dimensional Obsessive-Compulsive Scale
DIAMOND: Diagnostic Interview for Anxiety, Mood, and OCD and Related Neuropsychiatric Disorders
DASS-21: Depression, Anxiety, and Stress Scales

## Notes

### Competing Interest Statement

Jamie D. Feusner, Nicholas R. Farrell, Mia Nuñez, Nicholas Lume, Catherine W. MacDonald, Patrick B. McGrath, Larry Trusky, Stephen Smith, and Andreas Rhode report personal fees from NOCD Inc.

### Clinical Trial

NCT06466447

### Funding Statement

This analysis was funded by NOCD

### Author Declarations

The University of California Los Angeles IRB has determined the research to be exempt from IRB review and has stated the following: Based on the information that was provided in this application, this project does not meet the definition of Human Subject Research. Thus it is not necessary to submit the application to the IRB for review and approval.

### Summary of Updates

Table 1 expanded, Table 2 added, longitudinal follow-up results added, other text revisions.

